# Reducing SARS-CoV-2 infectious spreading patterns by removing S and R compartments from SIR model equation

**DOI:** 10.1101/2020.06.12.20127498

**Authors:** Charles Roberto Telles

## Abstract

This research points to the asymptotic instability of SIR model and its variants to predict the behavior of SARS-CoV-2 infection spreading patterns over the population and time aspects. Mainly for the “S” and “R” terms of the equation, the predictive results fail due to confounding environment of variables that sustain the virus contagion within population complex network basis of analysis. While “S” and “R” are not homologous data of analysis, thus with improper topological metrics used in many researches, these terms leads to the asymptotic feature of “I” term as the most stable point of analysis to achieve proper predictive methods. Having in its basis of formulation the policies adopted by countries, “I” therefore presents a stable fixed point orientation in order to be used as a predictive analysis of nearby future patterns of SARS-CoV-2 infection. New metrics using a Weinbull approach for “I” are presented and fixed point orientation (sensitivity of the method) are demonstrated empirically by worldwide statistical data.

## 1) INTRODUCTION

The nonlinear dimensionality reduction scenario pointed in this research refers mainly to the unpredictability of epidemiologic framework of SIR (susceptible, infected, removed) stochastic or deterministic models and its variants to track the possible rate of infection of the severe acute respiratory syndrome coronavirus 2 (SARS-CoV-2) among population.

The starting point to the limitation of predicting future rates of transmission was importantly and deeply noticed by Manzo and Roberts *et al* [1,2] during the pandemic spreading, and also briefly described by Merchant and other authors [3,4,5,6]. These authors presented descripted analysis of SIR models and variants failures to achieve a good to fit predictability and sensitivity of how SARS-CoV-2 infectious spreading patterns might occurs over time (also this limitation being live verifiable by automated tools data bases available to perform it at https://projects.fivethirtyeight.com/covid-forecasts/ [7] as exemplified in “S1 figure”. Many researches based on SIR models or its variants [8], just to mention very few examples [7,9-18] are confronted with barriers found mainly on the basis of S and R compartments of SIR model due to pandemic complex scenario involving societies and SARS-CoV-2 distinct spreading patterns of infection where those two terms of equations are now suspended. “S2 figure”

Despite new formulations based on agent-based modeling approaches [19,20] for SARS-CoV-2 are being deeply described [21-23], the lack of fixed point orientation and asymptotic instability for S and R are still visible when we consider that an agent-based model can be under or overestimated by the type and duration of policy making worldwide [24-33] having this feature as the main attractive network interactions. This confounding environment of study presents itself for COVID-19 pandemic as an impossible methodology to be used in terms of prediction since embedded components between policies and individual behavior are presented and act as a limiting factors for the machine learning analytics for S and R as well as the pre assumptions of a global homomorphism basis for the individual scale of infected person and SARS-CoV-2 behavior to population behavior.

Recent researches have been pointing to the robust analysis of convergence orientation within policies adoption by countries (non-pharmaceutical interventions (NPIs)) [24-33], therefore, remaining these efforts to the conclusion of component I of model as the best predictable pattern formation to be used for epidemics analytical behavior. Considering all countries epidemic behavior in convergence to the policies adopted, I component present a good performance of indicator of analysis behavior to infer with numerical results the noise [1-7] caused by some compartment components of SIR models approaches of predictability.

Even with high variance produced by other variables for S and R that influence the final solutions of the phenomenon, these fixed point stable parameters of I can create a confident region of statistical analysis [24-33] in terms of observation of the exponential growth of virus along days and therefore it could be more conclusive to many mathematical infectious disease models (SIR stochastic or deterministic approaches) that were created since the beginning of epidemics and later pandemics spreading.

Many variables that affect virus transmission rates [34], such as the type and duration of health policies adopted by each country (NPIs), health infrastructure, population genetics, human variance in terms of biological resistance (symptomatic, pre and asymptomatic cases), epidemic outbreak, infodemics worldwide (individual, community, government and media scale), globalization aspects, COVID-19 testing availability, reliability and time for results outcomes, virus mutation, and citizen adherence to social isolation and social distancing present a strong influence on the S and R behavior patterns. These confounding outcomes in each country pose a challenge to identify exact design and results of S and R patterns, therefore influencing on I compartment. However these inputs of data can create a confounding environment of study, where the mathematical simulation of SIR model finds limitations due to these non-homologous data [1-7] and consequently heteroscedasticity form for data results; in this sense, it is justifiable the analysis policies adopted by countries as the most reliable, at the moment [24-33], form of reducing COVID-19 cases while no vaccine or drugs present consistent and effective use for treating the disease or stop virus propagation, that could influence directly the S and R components, thus, performing a robust prediction analysis.

## 2) METHODOLOGY

### 2.1) The S and R constraints

In many phenomena, the level of randomness of a system assumes extreme asymptotic instability characteristics for variables convergence aspects as it is observed in this pandemic patterns [7]. The confounding environment of variables involved in the pandemic spreading patterns assume distinct values for the exponential behavior of infection over time not only for the same sample of analysis (country) [7], but samples compared to each other [35,36].

In order to extract fixed point orientation with not homologous data involved in the epidemics cases, concerning the system global homomorphism over time, no stability pattern was found for S and R in recent researches [7-23] except for I [24-33]. This stability feature for S and R is not commonly observed in the empirical terms of the analysis for short time periods of observation while these data presented time-varying empirical data and unresolved partial differential equations on the basis of epidemics evolution and this still applies for long time periods as well [1,2,7,37]. While S and R periodic fixed point that could uncover a stability pattern formation needed for a deterministic SIR modeling are not obtained (vaccines/drugs/immunization), results, nowadays [7-23] generates probabilistic distributions permeating the system for short time lengths (local homomorphism) and long time lengths (global homomorphism) generating distinct distributions shapes and scales for each country. These set of variables that make up the system assumes different behaviors over time periods and it is not possible to determine universally that the system have a persistent not embedding behavior in the way it was constantly observed in many researches using SIR and its variants models [7-23], having in the data sets the main limiting factor of analysis.

Now consider this premise that confounding or even spurious environment (not homologous) of variables presented by S and R compartments of SIR equation basis present high instability properties in terms of expressing convergent orientation of the global homomorphism for the infection spreading patterns, while I compartment of SIR model is asymptotic stable to the policies adopted by countries. For this point, the evolution of the disease spreading patterns need to be evaluated by the policies adopted by countries that present nowadays the most stable pattern of convergence [24-33] and therefore, most reliable and sensitive data can be used as tool for predictive analysis.

By excluding S and R compartments of equation and using only I variable as an indicator of infection spreading pattern, it exclude as well the limiting factors of SIR model of predictive analysis and therefore, the confounding environment of research. The compartment I resemble the growth of infection over days that depending on the policies adopted by a country it express higher or lower exponential growth behavior. If considering the exponential growth as how efficient are the policies adopted in order to reduce transmission patterns, then, we have an indicator of analysis that is pre assumed on fixed point orientation of infection spreading patterns, and hence, a valid indicator of countermeasures, monitoring aspects and expected results in a wide domain of policies to be adopted.

To achieve this indicator’s numerical results, we need to understand time periods of infection based on cumulative cases time series data limited to the exponential behavior of infection in its mean per population samples. The mean reflects the sensitivity needed to observe the phenomenon evolution over time and how stability patterns are occurring. For this point of observation, exponential growth mean resembles exactly how the infection spreading patterns have encountered adequate policy measures or inadequate fixed point orientation of the type of policy adopted as it will be explained in the next subsectio 2.2.

### 2.2 “I” of SIR model as the most reliable and sensitive indicator

For this analysis, it was considered the observed infected population samples *Y* = (*Y*_1_… *Y*_*n*_) of the general definition of the exponential behavior of infection as *f*(*Y* ; *λ*) = *λ e*^−*λ Y*^ where the samples are taken from zero cases until the observed maximum exponential growth reached per population amount for each country [35-36] under the *I*′ (new infection spreading patterns under policy measures influence) designed region of analysis.

In the shape behavior *k* = 1 or *k* > 1of exponential irregular distributions of SARS-CoV-2 infection defined in the original form as 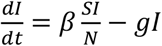, generically and instable [7-23] it can understood as 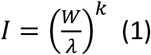, where the infected *I* is influenced by unpredictable scale of infection *λ* (*N*) with inconsistent behavior of variables transition rate (*βSI*) defined as *W*, and is not assumed for *gI* in the original form of R, that there are a normal distribution output for this virus spreading patterns, therefore it was used an unpredictable shape *k*, mainly defined those as components S and R of SIR model and its variants. This equation represents the presence of confounding variables environment *W* with unknown predictive scale of *exp λ* or maximum likelihood estimator for *λ* due to nonlinear inputs for S and R, and therefore generating nonlinear outputs *k* (asymptotic instability) [38] by the virus infection *I* over *Y* (population). The outputs with heteroscedasticity form for *k* and *λ* are removed of occurring as far as the *K* < 1 Weibull parameterization aspect [39] of distribution be elected as the most reliable region of analysis (attractive orientation), thus modifying the first equation (1) to 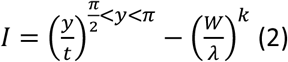, hence with the new SIR model proposition as *I* = *I*′ − *S* + *R*, where *I* is asymptotic to *I*′. This is a mandatory redesign since many scientific breakthroughs are pointing to policies as the best approaches to reduce COVID-19 nowadays [24-33].

This mean *y* indicated at (2) is found as a persistence diagram existence [40] by mapping each adjacent pair to the point (*f*(*Y*), *f*(*t*)) minimum and maximum observations, resulting in critical points of *y* function over time *t* in not adjacent form, thus expressing random critical values defined by 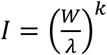 in the original form of observation of the event. But since we need to filter *f*(*Y*) − *f*(*t*) unstable critical points (oscillatory instability of S and R) to an attractive minimum behavior for the distribution functions *F* for S and R towards I, as 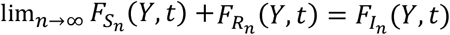, therefore, the redefinition of mean *y* over time is a necessary condition.

At this point, by rejecting the persistence diagram unstable critical points generated, a local minimum of the event as an average mean can be obtained by having *y* with the higher number of samples *Y* that finds a condition roughly described in the nonlinear oscillations within the exponential growth epidemic behavior of event as limited between local maximum growth defined by *π* by its half curvature oscillations 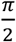 as a local minimum being non periodic (2 *π*) due to *κ* < 1. Therefore *y* assumes the desired oscillations samples and region condition like 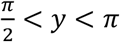 where persistent homology can be found for *t* ∴ *κ* < 1 to be situated in the oscillations pairing region of sin (*π*) = 1 and cos (*π*) = 0 for *y* desired coordinates (*f*(*Y*), *f*(*t*)) of stability with *t* + 1 as 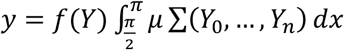, thus assuming the shape and limit to *κ* < 1. Then concerning time lengths of samples, it is designed as *t*(*x* + 1) … *f*(*Y*)*μ* ∑ (*Y*_0_… *Y*_*n*_) *dx* starting from *t*_0_, …, *t*_*n*_ ≤ sin(*y*) results in the desired data distribution with a conditional shape of Weibull parameterization *κ* < 1 for the analysis, therefore rejecting any critical value beyond cos (*π*) = 0 and under sin (*π*) = 1.

In this simple and overall form, redefining the mean shape for the infection spreading patterns of *I*′ approximately as *y* = 1/*λ* under *κ* < 1, the numerical representation of the scale considering a global homomorphism of the persistent homology [40] observed can be limited by the highest number of scales that fitted best by the observed exponential mean *Y* scale of *I*′ until it reaches a desired form like *y* = *Y*.

This modeling is represented at figure 1 for United States analysis on 02, Jun, 2020 [35]. At this point the days counting forward this condition will be rejected to extract the exponential infection spreading rate in the formula *rate* (*R*) = *y*/*t* and *t* = *κ* only in the desired event expression. This approach can be more sensitive in terms of time overview of the disease and its potential to infect as time passes. This sensitivity is much more confident for predictions due to the exponential behavior of infections at community phases of infection spreading patterns.

**Figure 1.**
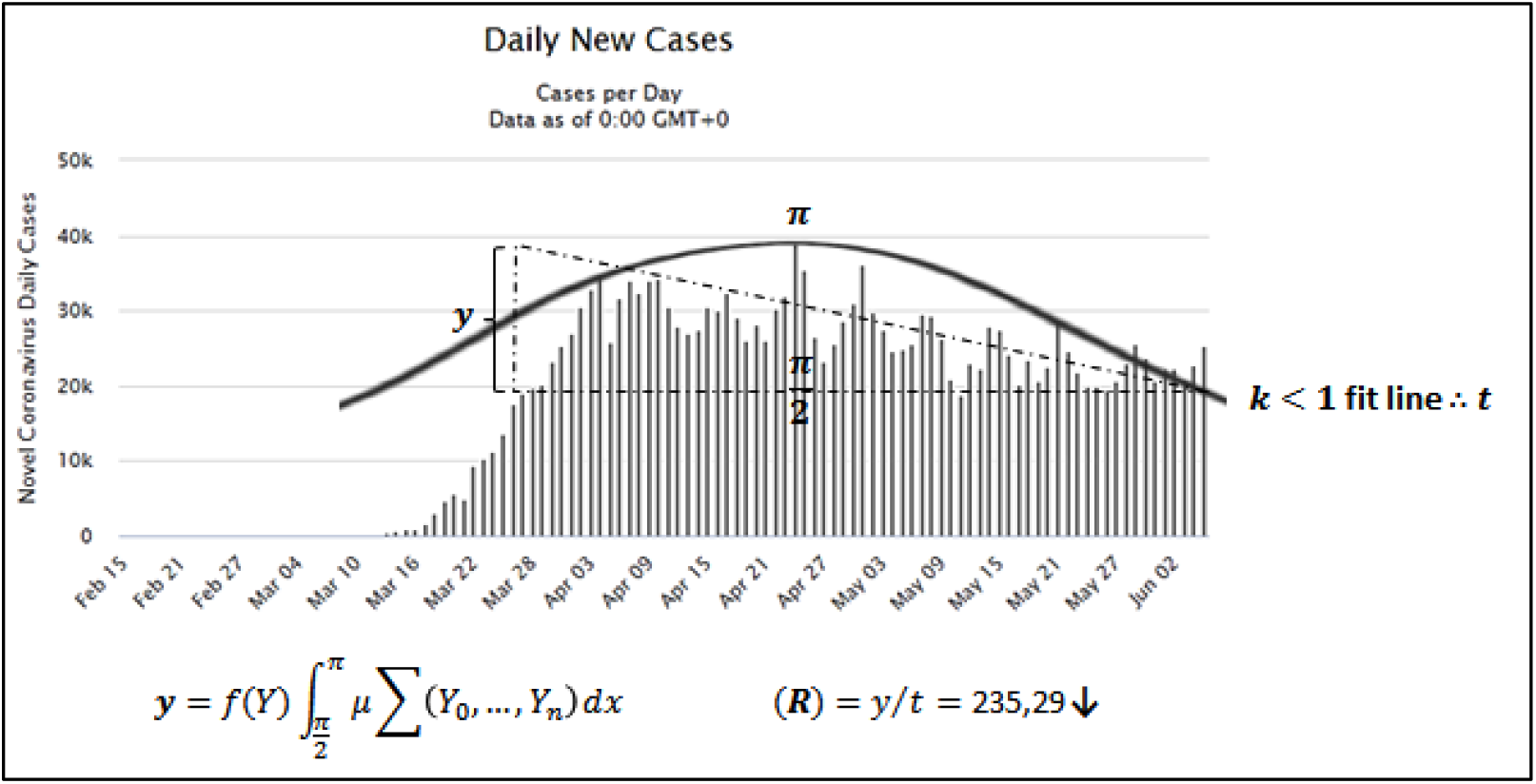
Exponential infection spreading rate for United States. (*R*) analysis obtained for 02 Jun, 2020. Note that persistent homology is now showed in the figure. However it can be induced by hypotenuse angle proximity. Data source: Worldometer.

Now considering the critical points of *I* found per population *y* margins that infection has reached, it is counted as *I*′ the positive exponential growth in the mean *y* found at the region limited to 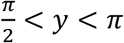 for first, second or other waves of infection, counting for this second or other period with cumulative time since outbreak (but not cumulative population ratio found for the first wave of infection or the previous ones).

At Table 1, for the second column, it presents the growth of infection (minimum values) per amount of population *y* observed for a given sample (country) by the empirical observations, therefore, in the proposition *I* = *I*′ − *S* + *R*, it gives the values of the exponential growth mean reached in the *I*′ shape in average days peaks of infection since the outbreak until the condition *κ* < 1 and therefore not counting as a mean *y* higher values than the ones found at *I*′ − *S* + *R*. The third column “*t*” presents how much days the infection presented an exponential growth with a mean reached. And finally the fourth column (*R* approach have similarities with SIR models, but it is based on distinct aspects of analysis of variables S and R by removing these variables from the formula and focusing mainly in the I variable defined by Weibull parameterizations and exponential desired distributions. This design of analysis is very relevant due to instability aspects of SIR analysis taken since outbreak of disease, mainly existing in the S and R variables due infodemics, type of analysis conducted for this research (the apparent lack of overall topological homology of data) and other nonlinear aspects of the new coronavirus 2 disease. For this reason, the proposed method of analysis considers only the infectious disease aspect of evolution of cases rather than pre assuming full immunity or deterministic models for population behavior that is in this case one of the most influencing forms of keeping the virus active in its propagation.

**Table 1.** COVID-19 exponential growing patterns per population and time period by country/region.

| Country | $y$ | $t$ | $R$ on 01 May |
| --- | --- | --- | --- |
| Worldwide | 60.000 | 99 | 606,06↓ |
| Europe | 30.000 | 91 | 329,67↓ |
| Italy | 4.000 | 51 | 78,43= |
| South Korea | 250 | 20 | 12,5= |
| China | 1.500 | 28 | 53,57= |
| Iran | 2.000 | 46 | 43,47= |
| Spain | 5.000 | 55 | 90,90↓ |
| France | 4.000 | 45 | 88,88↓ |
| United States | 30.000 | 53 | 566,03↑ |
| Brazil | 5.000 | 51 | 98,03↑ |
| Germany | 4.000 | 41 | 97,56↓ |
| Russia | 5.000 | 46 | 108,69↑ |
| United Kingdom | 5.000 | 58 | 86,20↑ |
| Singapore | 500 | 57 | 8,77↑ |
| Portugal | 500 | 49 | 10,20↓ |
| India | 1.000 | 58 | 17,24↑ |
| Canada | 1.500 | 48 | 31,25↑ |
| Japan | 500 | 67 | 7,46↑ |
| Sweden | 500 | 58 | 8,62↑ |
| Argentina | 100 | 57 | 1,75↓ |
| Chile | 500 | 52 | 9,61↑ |
| Saudi Arabia | 1.000 | 54 | 18,51↑ |
| United Arab Emirates | 400 | 70 | 5,71↓ |
| Egypt | 200 | 62 | 3,22↑ |
| Pakistan | 500 | 53 | 9,43↑ |

The $y$ and $t$ data are retrieved for 01 May, 2020. Data source: Worldometer.

What are the main frontiers to achieve a stability behavior concerning data science and big data in order to build a robust performance for predictive and behavior analytics of complex events of COVID-19 spreading patterns of infection? Based on this introductory statement we have the following points to address:

### a. Micro-processes under a given assumed stability

That in this case assumes a monotonically increasing exponential behavior for I component even with the not homologous data presented in the complex event as S and R and therefore the indicator is limited directly to the policies adopted by countries as the main stability point of analysis for *κ* < 1. At this point, the micro-processes for S and R are the main constraint for the SIR model traditional application in the case of SARS-CoV-2 modeling equation.

### b. Time and machine learning/mathematical simulations experience

Defined as a local homomorphism of solutions for S and R, the SIR predictions performed until now fail to assume a global homomorphism for all epidemics duration and its behavior for each sample (country) analyzed. This is caused mainly due to observations of short time lengths of the infection event in which possibly the confounding variable’s environment of S and R aren’t the kind of data in empirical terms that gives origin to recurrent plots or defined phases spaces, and also not being possible to apply this same results to other sample in terms of size, time and spreading patterns. Considering I as the unique feature, stable analysis and phase space can be extracted among samples and mainly having all those comparisons based on the same root of observation parameters, a feature that makes the analysis robust and predictive as far as observations are taken from empirical data at specific shape and scale distribution of the event.

### c. Empirical observations and data constraint (the lack of persistent homology)

To perform a reliable analysis on such set of data for S and R one would have to correct for multiple factors for each country, such as phase of the epidemic, time from the virus introduction into population to start of mass testing campaigns, case definition, number of tests performed per million population, sensitivity of tests used, testing criteria, population compliance, testing availability in different regions of a country, estimated ratio of detected and undetected cases, etc. This confounding environment of analysis for each country assume high dimensional order and nonlinear oscillations together with item 1, 2 and 4 due to complex scenario each country has in its social, economic, political, scientific and cultural aspects and are considered as the main cause for S and R failures rates of predictive analysis of SARS-CoV-2 with SIR deterministic or stochastic models variants.

### d. Privacy and internet of things

Concerning mobile apps and person-to-person surveys to detect SARS-CoV-2 features, many statistical results present possibly false information, subjectivity of data and limited scope of analysis due to ethical or policies implications. For these limiting points the data sets used to estimate values for S and R can challenge serious flaws and inconsistency. These factors sum up to the item 3 constraints and it was not addressed further in this research, however this was considered as a consistent point of constraint for S and R data retrieved worldwide [41,42].

Based on a local homomorphism existent for *κ* < 1 shape based on (*R*) = *y*/*t* scale of distribution, a consistent fixed point orientation can be observed in these results section, therefore confirming the use of the indicator as a predictive tool designed for policies measures adopted to reduce COVID-19 infection spreading patterns.

## 3) RESULTS

### 3.1 A predictive tool for policy measures

How can governments precisely state “let’s keep the policies as they are or let’s adopt new ones or change the old ones scope or criteria?”. This answer also addresses to the question: how much sensitive of data retrieved and fixed point orientation have the policies adopted among countries for predictive analysis?

Now after explaining the methodology used, Table 2 presents, from the epidemics outbreaks until days considered as 7 and 13 April, 7 days were the amount of time is taken and the behavior of infection obtained for prediction and sensitivity, indicated by arrows, can be with high confidence in order to make predictions as it can be observed by differences in the nearby future. In the 25 samples analyzed, 12 presented (orange color) different behavior of infection spreading patterns while 13 expressed a constant evolution of infection that indicates also a positive analysis for predictive statistics even with low amount of time considered (7 days sensitivity and prediction).

**Table 2.** Sensitivity of time and exponential spreading patterns confronted with policy measures adopted by countries.

| Country | R on 7 April | R on 13 April |
| --- | --- | --- |
| Worldwide | 675,67↑ | 886,07↑ |
| Europe | 410,95↑ | 379,74↓ |
| Italy | 90,90↓ | 78,43↓ |
| South Korea | 12,5↓ | 12,5= |
| China | 53,57↓ | 53,57= |
| Iran | 44,44↑ | 43,47↓ |
| Spain | 131,57↑ | 121,95↓ |
| France | 138,88↑ | 100↓ |
| United States | 625↑ | 540,54↓ |
| Brazil | 25↑ | 28,57↑ |
| Germany | 105,26↑ | 100↓ |
| Russia | 15,62↑ | 27,02↑ |
| United Kingdom | 64,51↑ | 69,64↑ |
| Singapore | 0,96↑ | 1,75↑ |
| Portugal | 15,15↑ | 13,15↓ |
| India | 8,33↑ | 9,43↑ |
| Canada | 27,77↑ | 24,39↓ |
| Japan | 3,92↑ | 7,14↑ |
| Sweden | 5,71↑ | 0,65↓ |
| Argentina | 1,51↑ | 2,56↑ |
| Chile | 10↑ | 8,33↓ |
| Saudi Arabia | 3↑ | 4,16↑ |
| United Arab Emirates | 4,34↑ | 5,76↑ |
| Egypt | 1,31↑ | 2,27↑ |
| Pakistan | 3,44↑ | 5,71↑ |

Data source: Worldometer.

While in 7 days of predicting analysis, several countries implemented new policies [43,44] measures to reduce SARS-CoV-2 spreading patterns and prediction was affected highly by this measures. Note that no specification of policy measures was provided. The objective of this research is not to determine which measure gives the best result, but just to provide the observation that policies do have influence on the COVID-19 spreading patterns. For further interests on the type, duration and efficacy of measures adopted by countries, refer to [24-33]. Table 3 presents, from the epidemics outbreaks until days considered as 13 April to 01 May, counting for higher time period of analysis. Only 7 days more were added on 13 April results since 07 April when policy measures were vastly started or in course among countries. For this new predictive results based on policies influences adopted, for 19 days in future, the behavior of infection obtained for prediction on 13 May, indicated by arrows, were modified on 01 May. In this period, the same as happen before, new policies measures were adopted or in course by countries [43,44], thus influencing the predictive/sensitivity analysis. In the 25 samples analyzed, 9 presented (orange color) different behavior of infection spreading patterns while 16 expressed a constant evolution of infection that indicates also a positive analysis for predictive statistics even with low amount of time considered (19 days sensitivity and prediction).

**Table 3.** Sensitivity of time and exponential spreading patterns confronted with policy measures adopted by countries.

| Country | *R* on 13 April | *R* on 01 May |
| --- | --- | --- |
| Worldwide | 886,07↑ | 606,06↓ |
| Europe | 379,74↓ | 329,67↓ |
| Italy | 78,43↓ | 78,43= |
| South Korea | 12,5= | 12,5= |
| China | 53,57= | 53,57= |
| Iran | 43,47↓ | 43,47= |
| Spain | 121,95↓ | 90,90↓ |
| France | 100↓ | 88,88↓ |
| United States | 540,54↓ | 566,03↑ |
| Brazil | 28,57↑ | 98,03↑ |
| Germany | 100↓ | 97,56↓ |
| Russia | 27,02↑ | 108,69↑ |
| United Kingdom | 69,64↑ | 86,20↑ |
| Singapore | 1,75↑ | 8,77↑ |
| Portugal | 13,15↓ | 10,20↓ |
| India | 9,43↑ | 17,24↑ |
| Canada | 24,39↓ | 31,25↑ |
| Japan | 7,14↑ | 7,46↑ |
| Sweden | 0,65↓ | 8,62↑ |
| Argentina | 2,56↑ | 1,75↓ |
| Chile | 8,33↓ | 9,61↑ |
| Saudi Arabia | 4,16↑ | 18,51↑ |
| United Arab Emirates | 5,76↑ | 5,71↓ |
| Egypt | 2,27↑ | 3,22↑ |
| Pakistan | 5,71↑ | 9,43↑ |

Data source: Worldometer.

It is possible to observe that countries who didn’t change its policies measures [43,44] didn’t express modifications obtained by this predictive method. This happened even for countries who already understood its data as a good fit to reduce SARS-CoV-2 spreading patterns as well as countries that didn’t make any modification due to countries personal reasons.

Now from epidemics outbreaks until 02 June that gives 33 more days of the event evolution at Table 4 having as parameter the data of 01 May, even with oscillatory dynamics of policies adopted, one might assume that as far as policies don’t change anymore, the predictive method will point to a very confident method to predict nearby future analysis of infection spreading patterns. For this concern, since sensitivity is very connected to policies, the predictive power can be evaluated for any given future time of analysis by it. In this form, this research states that no matter how S and R will affect the event, the convergent attraction of policies can provide robust analysis of infection spreading patterns in terms of prediction and sensitivity.

**Table 4.** Sensitivity of time and exponential spreading patterns confronted with policy measures adopted by countries.

| Country | R on 01 May | R on 02 June |
| --- | --- | --- |
| Worldwide | 606,06↓ | 572,51↓ |
| Europe | 329,67↓ | 162,6↓ |
| Italy | 78,43= | 78,43= |
| South Korea | 12,5= | 0,09↓ |
| China | 53,57= | 53,57= |
| Iran | 43,47= | 25,64↓ |
| Spain | 90,90↓ | 90,90= |
| France | 88,88↓ | 3,89↓ |
| United States | 566,03↑ | 235,29↓ |
| Brazil | 98,03↑ | 180,72↑ |
| Germany | 97,56↓ | 2,73↓ |
| Russia | 108,69↑ | 96,15↓ |
| United Kingdom | 86,20↑ | 22,22↓ |
| Singapore | 8,77↑ | 5,61↓ |
| Portugal | 10,20↓ | 1,85↓ |
| India | 17,24↑ | 64,44↑ |
| Canada | 31,25↑ | 9,37↓ |
| Japan | 7,46↑ | 0,30↓ |
| Sweden | 8,62↑ | 6,66↓ |
| Argentina | 1,75↓ | 6,17↑ |
| Chile | 9,61↑ | 41,66↑ |
| Saudi Arabia | 18,51↑ | 17,44↓ |
| United Arab Emirates | 5,71↓ | 5,39↓ |
| Egypt | 3,22↑ | 7,44↑ |
| Pakistan | 9,43↑ | 23,52↑ |

Data source: Worldometer.

Now from 01 May to 02 June, in the 25 samples analyzed, 11 presented different behavior (orange color) of infection spreading patterns while 14 expressed a constant evolution of infection that indicates also a positive analysis for predictive statistics even with high amount of time considered (33 days sensitivity and prediction).

By performing these examples of analysis, it can be observed that prediction is highly associated with the policies adopted by countries, and therefore the predictive power is intrinsically associated with the sensitivity power of analysis over time.

One important observation of the method proposed given by the author due to his nationality, is about Brazil COVID-19 spreading patterns where since beginning of local epidemics, each state adopted distinct measures without a federal whole pack of actions to be performed for public policies (health and economics). This initial divergence among states provided the infection spreading patterns to be in a heteroscedasticity form for statistical descriptive data. The false feeling that some states were more infected than others and mainly empirical based methods used to start policies interventions made the exponential power of infection grow beyond any type of scientific study conducted to predict the infection spreading patterns. This was mainly caused by the observations of this research, due to S and R parameters of analysis and constraints. Also, if observing data of tables 1-4 for Brazil, you are going to see that policies used by country as a whole didn’t find any other instability aspect for COVID-19 reduction. These results for the country Brazil, resembles exactly what happened here along the time: no default policies for the whole country, several changes in policies adopted by states individually (trying to find what gives the best results), no use or belief in scientific data and very poor population compliance/confidence to measures randomly taken.

That’s the author analysis of his country of origin based on political scenario of country and indicator metrics obtained.

## 4) DISCUSSION

It is not needed that all countries present the same date of implementing measures to prove the article findings since distinct phases of epidemics among countries and regarding policies measures starting date are not a deterministic model in real life, but both data present high asymptotic stability, therefore, being predictable and observable by the proposed mathematical modeling.

Another point for removing S and R terms of SIR model equation is related to table’s data and random dates that refer to different epidemic phases of data collection for each country. These distinct phases are important to be considered together in any sample collection due to need of a methodology that can extract behavior of the disease in the not optimal evolution (SIR deterministic approaches) of the virus infection and policies adopted by countries, hence, revealing in the complex scenario the disease dynamics under a confounding environment of data with more sensitivity and possible rapid response of countries policies. The homology line of research conducted here applies only for the first exponential maximum increase of the phenomena and it can be tracked topologically by the policies adopted by each country over time, even excluding any other confounding variable existent in the event, since it will not affect the overall convergence of the phenomena.

All the variables that influence COVID-19 spreading patterns is exactly what this article points to be observed as they are confounding and it is needed to extract an indicator that can point to the high asymptotic effect of the phenomena, that is in this case the policies adopted. The empirical results from China and South Korea are the main evidence on it. Considering it there is not reason to correct these confounding factors with numerical results in the topological field by now since we have strong indicative that all this random data influence can be asymptotically unstable (not convergent) to the policy measures.

Following these statements, and by these data being not homologous, the analysis for this relations lead to high degree of heteroscedasticity that can be observed not in terms of variance analysis or linear regression analysis but a general overview of distribution shapes and scales with Weibull parameterization in order to analyze only the exponential behavior related to the formation of normal distributions phase or tendencies at marginal measures. In this way, both homologous and not homologous topologies are pre assumed qualitatively (QDE) for a preexistence of fixed point orientation of the event. In this way, the method proposed reveals patterns of homology formation in the invisible condition of detecting each variable’s coupling behavior effects.

The homology is obtained by considering as stated in the Weibull approach by excluding any data related to the minimum value of the exponential curve before a second wave is observed. One other advantage of the method is that there is no need to describe the event in its full behavior (very long time lengths) as well as wait for longer time series to be able to make predictions about how policies influence the event. This was not proven by numerical results on the algebraic topology field, but it was presented by the numerical results towards the probabilistic distributions observed in the data that reflects, among other countries, based on China and South Korea margins, the exact point of homology among the confounding variables from the beginning of the outbreak until the mean point of the exponential behavior of the event over time.

## 5) CONCLUSION

A method to be used as qualitative indicator of epidemics spreading patterns is offered and it is based on how policy measures influence the growing of exponential curve. Formula, results and discussion of topological framework is briefly presented in order to sustain the robustness of mathematical and empirical framework of analysis. This approach is mainly addressed to government’s interventions on public health measures against COVID-19 and also for researches to analyze independently for your own country how policies affected the evolution of virus spreading patterns.

The results obtained showed that the indicator can be valid as a predictive method of analysis in terms of identifying how the epidemics are behaving over time being influenced by public health policies. The method used is high sensible to policies adopted by countries having a confidence power relative to how a country respond to epidemic in terms of these policies.

Strategic policy interventions can be proposed while events takes place over time as well as evaluations of how policies already adopted can be failing to achieve the desired results. This feature can be used both for country or regions/states/localities within a country. Policy measures are considered nowadays as the best optimal solution to contain COVID-19 spreading patterns towards exponential growing aspect.

The COVID-19 event was considered from a theoretical viewpoint using the qualitative theory of differential equations (QED) framework to help to understand how the input of many variables and output results in terms of convergence and stability aspects of policies adopted by each country, could point to any possible confounding data related to empirical evidences of countries divergences visible through cumulative daily new cases and time for an exponential growing behavior of infection.

The authors also invite/suggests other researchers to perform the same analysis for their own country in other publications. Observe in details how indicator behaves and at what time (date) policies were adopted, as well as other aspects such as integrity of the measure, regions that adopted it, population compliance, duration and other strong variables influence like economics aspects. A machine learning tool could be used to track these results as an informative graph per country/region/city/public area/local building with specific range of time crossing the data of policy adopted/modified by one week, two weeks, one month and finally three months. That could be a good start to help governments or other interested people to retrieve predictive analysis and strategic plans to achieve COVID-19 spreading patterns reduction.

## Data Availability

All data are available on the manuscript and Worldometer website.

https://www.worldometers.info/coronavirus/

## Acknowledgments

I would like to thank all journals that have made COVID-19 research free to read, responding the call to action from the Office of Science and Technology Policy (OSTP) and other governments. This policy action was vital for researchers around the world to publish new discoveries about the virus.

## Author Contributions

Not applicable.

## Statement of Ethics

No humans or animals were involved in this study. Ethics approval was not required.

**No trial registration is applied for this study**

## Data availability

a. All data about COVID-19 daily cases over time are held in a public repository: https://www.worldometers.info/coronavirus/.
b. All data about COVID-19 policies adopted by countries are held in a public repository:
  1. https://www.ilo.org/global/topics/coronavirus/country-responses/lang--en/index.htm
  2. https://www.imf.org/en/Topics/imf-and-covid19/Policy-Responses-to-COVID-19#A
c. All other relevant data are within the manuscript and its Supporting Information files.

## Supplementary Materials

**“S1 Figure”.**
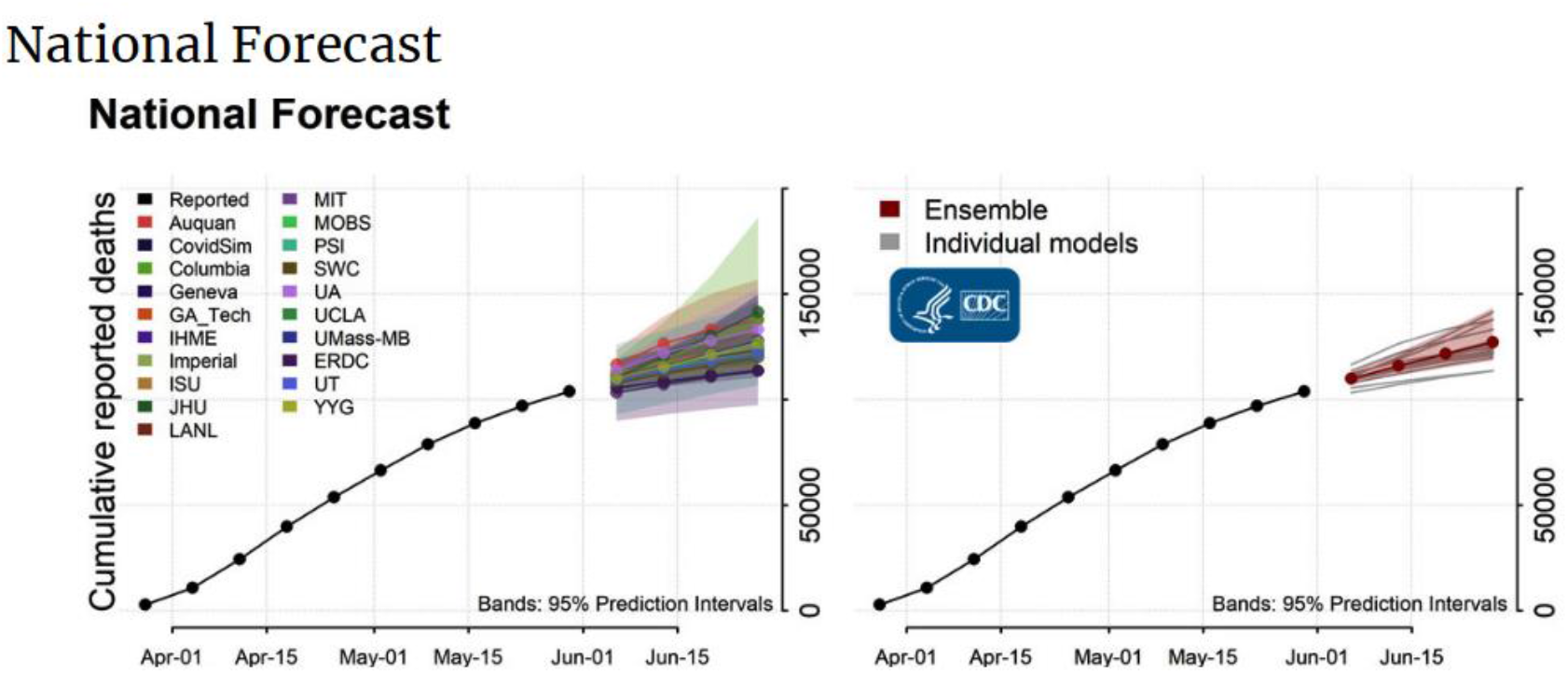
**Title:** Uncertainty of predictive methods with automated tools. Source: Centers for Disease Control and Prevention (CDC). Retrieved from: https://www.cdc.gov/coronavirus/2019-ncov/covid-data/forecasting-us.html, on June 11, 2020. **Legend:** Mostly of the models present a shaded region where prediction might face uncertainty degree of confidence. S1 supporting information image from CDC is under license of Public Health Image Library (PHIL): royalty-free.

**“S2 Figure”.**
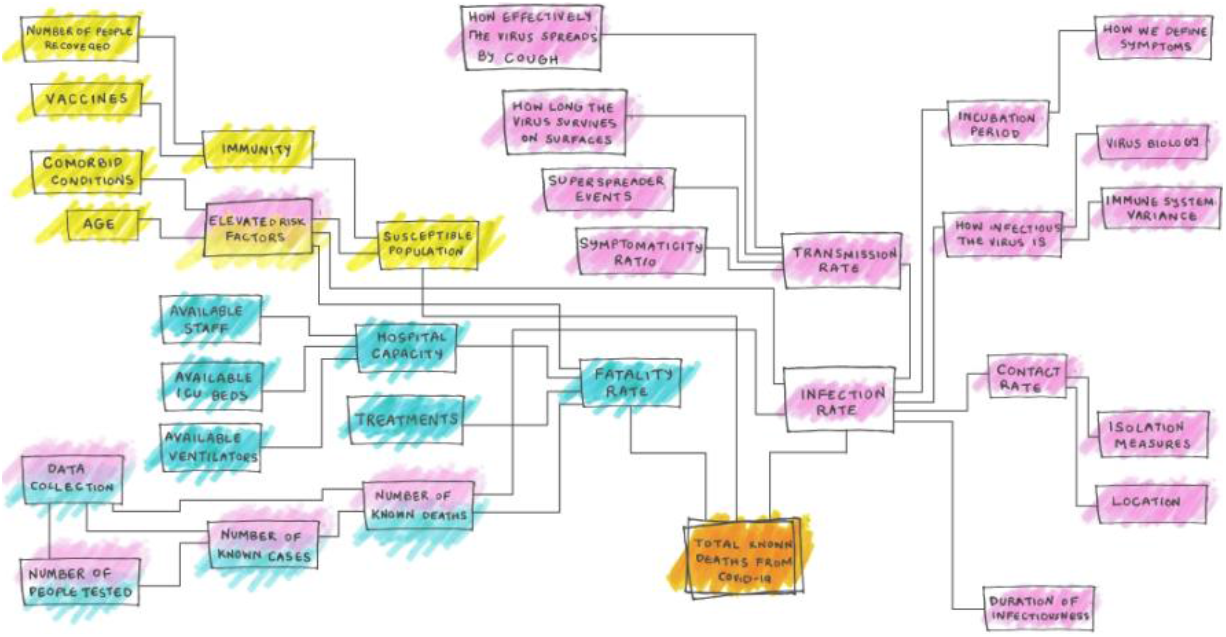
**Title:** Confounding environment of variables that influence COVID-19 infectious spreading patterns. **Image rights - Authors:** By Maggie Koerth, Laura Bronner and Jasmine Mithani. **Title of featured article:** “Filed under Coronavirus”. **Published** Mar. 31, 2020. **Source:** ABC News Internet Ventures. All rights reserved – Five Thirty Eight. **Retrieved from:** https://fivethirtyeight.com/features/why-its-so-freaking-hard-to-make-a-good-covid-19-model/, on June 11, 2020. S2 supporting information image from ABC News Internet Ventures. All rights reserved – Five Thirty Eight and the author have received permission to reuse as far as it has the proper attribution.

